# COVID-19 Epidemic Analysis using Machine Learning and Deep Learning Algorithms

**DOI:** 10.1101/2020.04.08.20057679

**Authors:** Narinder Singh Punn, Sanjay Kumar Sonbhadra, Sonali Agarwal

## Abstract

The catastrophic outbreak of Severe Acute Respiratory Syndrome - Coronavirus (SARS-CoV-2) also known as COVID-2019 has brought the worldwide threat to the living society. The whole world is putting incredible efforts to fight against the spread of this deadly disease in terms of infrastructure, finance, data sources, protective gears, life-risk treatments and several other resources. The artificial intelligence researchers are focusing their expertise knowledge to develop mathematical models for analyzing this epidemic situation using nationwide shared data. To contribute towards the well-being of living society, this article proposes to utilize the machine learning and deep learning models with the aim for understanding its everyday exponential behaviour along with the prediction of future reachability of the COVID-2019 across the nations by utilizing the real-time information from the Johns Hopkins dashboard.

## 1 Introduction

Coronaviruses are a large family of viruses that can cause severe illness to the human being. The first known severe epidemic is Severe Acute Respiratory Syndrome (SARS) occurred in 2003, whereas the second outbreak of severe illness began in 2012 in Saudi Arabia with the Middle East Respiratory Syndrome (MERS). The current outbreak of illness due to coronavirus is reported in late December 2019. This new virus is very contagious and has quickly spread globally. On January 30, 2020, the World Health Organization (WHO) declared this outbreak a Public Health Emergency of International Concern (PHEIC) as it had spread to 18 countries. On Feb 11, 2020, WHO named this “COVID-19”. On March 11, as the number of COVID-19 cases has increased thirteen times apart from China with more than 118,000 cases in 114 countries and over 4,000 deaths, WHO declared this a pandemic.

As the outbreak of the COVID-19 has become a worldwide pandemic, the real-time analyses of epidemiological data are needed to prepare the society with better action plans against the disease. Since the birth of novel COVID-19 [1], the world is restlessly fighting with its cause. As of April 1, 2020, based on the globally shared live data by the Johns Hopkins dashboard, worldwide there are 932,605 confirmed cases, out of which 193,177 are recovered and 46,809 lost their lives [2]. COVID-19 belongs to the family of the SARS-CoV and MERS-CoV, where it begins with the initial level symptoms of the common cold to severe level of respiratory diseases causing difficulty in breathing, tiredness, fever, and dry cough [3]. Prasad et al. [4] observed that the identification of the virus can be improved by imaging using immunoelectron microscopy techniques [5]. Figure 1 shows the typical structure of COVID-19 virus, from the throat swab of the first Indian laboratory-confirmed case, captured using sample electron microscopy imaging. Till date, detailed morphology and ultrastructure of this virus remain incompletely understood, and there are no specific vaccines or treatments for COVID-19. However, many ongoing clinical trials are evaluating potential treatments.

**Fig. 1.**
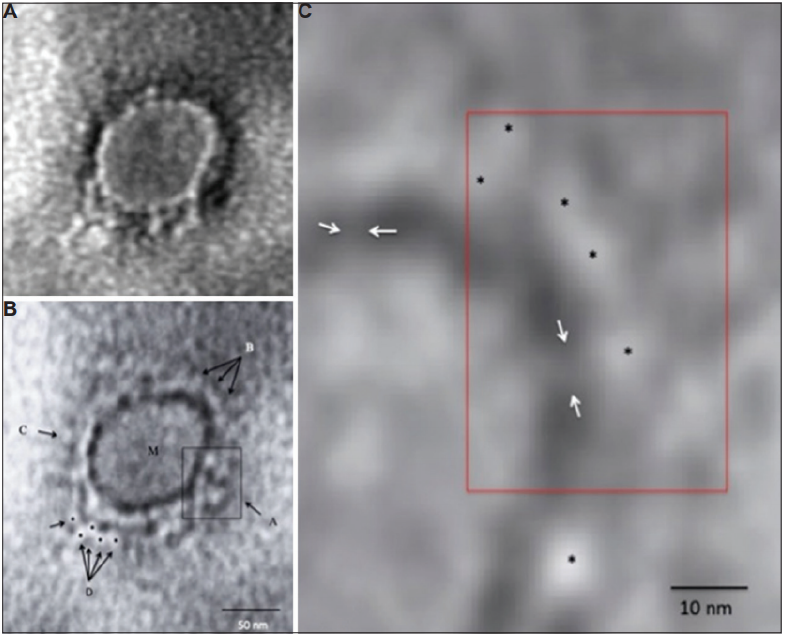
Electron microscopy imaging of COVID-19 representing morphological features at varying magnification levels [4].

### 1.1 COVID-19 transmission stages

As per the WHO reports [5], the pandemic situation is classified into four stages. The first stage begins with the cases reported for the people who travelled in already affected regions, whereas in the second stage, cases are reported locally among family, friends and others who came into contact with the person arriving from the affected regions. At this point the affected people are traceable. Later, the third stage makes the situation even worse as the transmission source becomes untraceable and spreads across the individuals who neither have any travel history nor came into contact with the affected person. This situation demands immediate lockdown across the nation to reduce the social contacts among individuals and control the rate of transmission. Figure 2, presents the outbreak of COVID-19 across the nations with the high number of confirmed cases such as US, Italy, Spain, and China based on the WHO reports [3], whereas figure 3 illustrates the rising characteristic of the number of confirmed, death, and recovered cases in these nations between the period January 22, 2020, to April 1, 2020, taken from Johns Hopkins live dashboard. The worst of all, stage 4 beings when the transmission becomes endemic and uncontrollable. Figure 4 presents the complete stages of the COVID-19 epidemic. Until now, several countries have entered stage 4. China is the first nation that experienced the stage 4 of the COVID-19 transmission. Though, it is claimed that the origin of the virus is Wuhan, China [6]; it affects the other developed countries (USA, Italy, Spain, Britain, etc.). These countries are now in stage 4 of the transmission and facing more number of infections and deaths compared to China. In the case of China, it is observed that exponential growth of the confirmed cases reaches the saturation stage where the number of cases stopped growing. This follows from the fact that the number of susceptible people, which are exposed to virus, are dramatically reduced. This was made possible due to the reduced social contact among people by segregating the infected individuals in quarantine and a complete lockdown period was initiated by the Chinese government, thereby reducing the possibility of further spread.

**Fig. 2.**
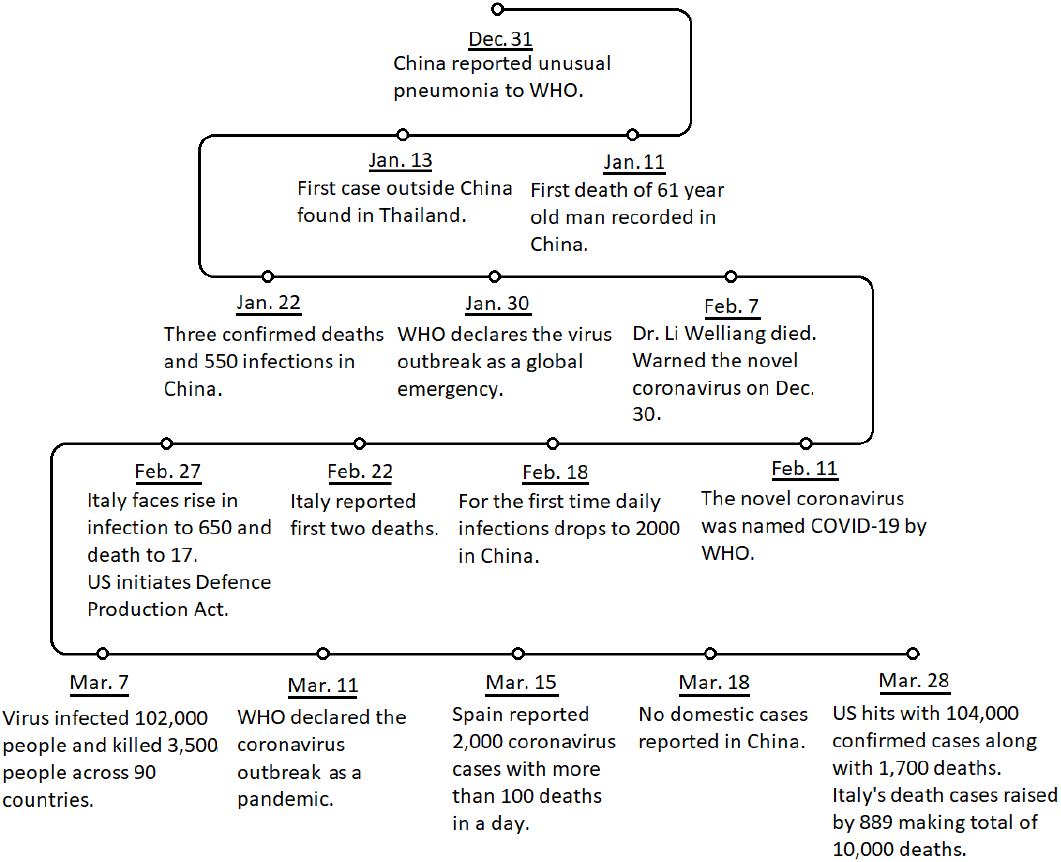
Timeline of COVID-19 across the nations.

**Fig. 3.**
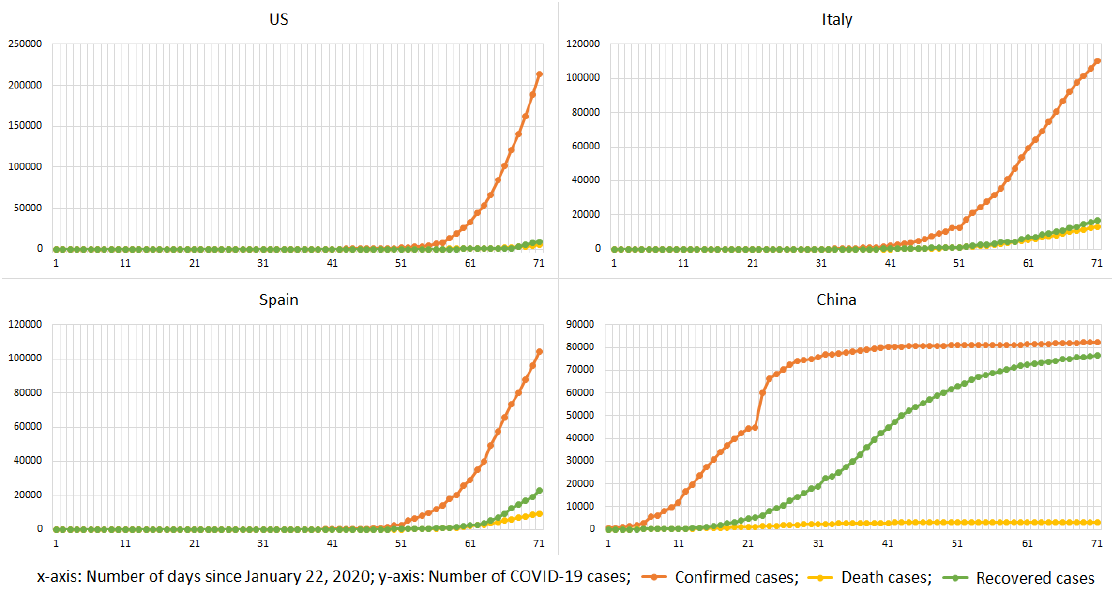
Epidemic status of the COVID-19 across nations having the highest number of confirmed cases since January 22, 2020.

**Fig. 4.**
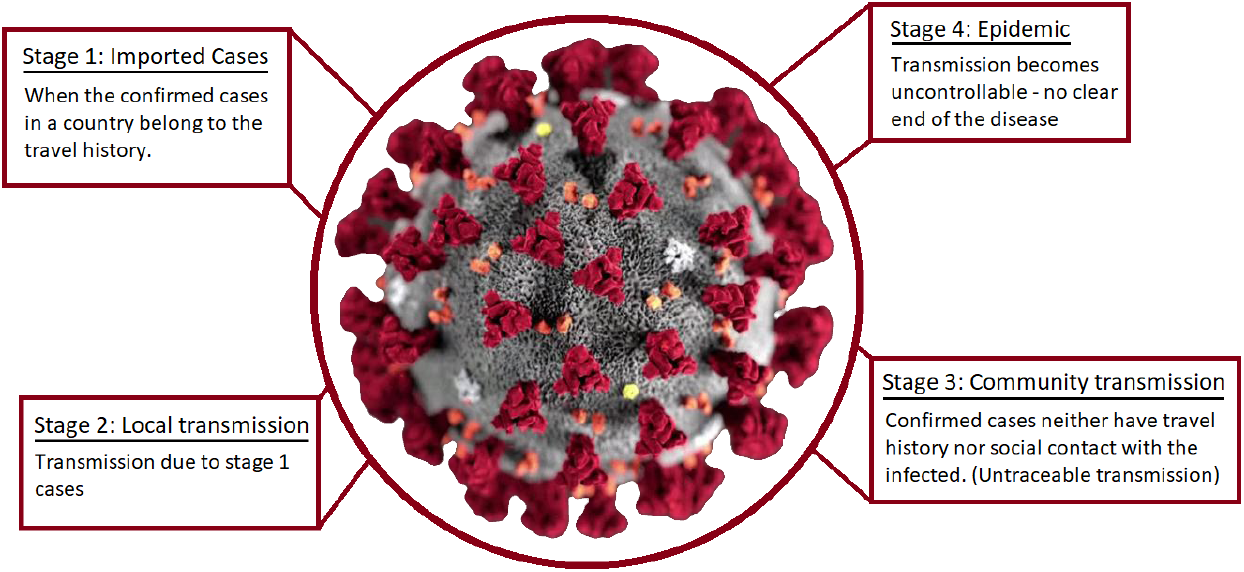
COVID-19: complete transmission stages.

Machine learning algorithms play an important role in epidemic analysis and forecasting [7], [8]. In the presence of massive epidemic data, the machine learning techniques help to find the epidemic patterns so that the early action can be planned to stop the spread of the virus. In this research, machine learning and deep learning models are used to observe everyday behaviour along with the prediction of future reachability of the COVID-2019 across the nation by utilizing the real-time information from the Johns Hopkins dashboard.

Rest of the paper is organised as follows: section 2 contains a brief discussion on existing machine learning based approaches, whereas section 3 talks about the dataset used for experiments. Epidemic analysis and experimental strategy are discussed in section 4 and 5 respectively, whereas results are discussed in section 6. Concluding remarks and future work are discussed in section 6.

## 2 Related work

Since the last decade, digital technologies are playing critical roles in major health sector problems including disease prevention, the present worldwide health emergency also seeking technological support to tackle COVID-2019. In a paper, authors highlighted the possible applications of trending digital technologies such as internet of things (IoT), big-data analytics, artificial intelligence (AI), deep learning and blockchain technology to develop strategies for monitoring, detection and prevention of epidemic; and also to identify the impact of the epidemic to the healthcare sector [9]. In a research work proposed by Benvenuto et al. [10], authors proposed an autoregressive integrated moving average (ARIMA) model to predict the spread of COVID-2019. In this paper, the author forecasted the various parameters for the next 2 days based on the study about the prevalence and incidence of the COVID-2019. This research work also demonstrates the correlogram and ARIMA forecast graph for the epidemic incidence and prevalence.

Deb et al. [11] proposed a time series method to analyze incidence pattern and the estimated reproduction number of COVID-19 outbreak. They performed statistical analysis to explore the trends of the outbreak to highlight the present epidemiological stage of a region so that various policies can be identified to address COVID-19 pandemic in different countries cite11. As per the present situation, it is essential to understand the early spread patterns of the infection to plan and control the effective safety measures. In this direction, Kucharski et al. proposed a scientific model of critical SARS-CoV-2 transmission by using different datasets to study the COVID-19 outbreak inside and outside Wuhan. With this, they explored the possible spread of disease outbreak outside Wuhan [12].

Recently there are several studies conducted on the epidemiological outbreak of COVID-19 using exploratory data analysis (EDA) based on various available datasets. The studies mainly focus on the occurrence of confirmed, death, and recovered cases in Wuhan and the rest of the world to understand the suspected threats and subsequent planning of containment activities [13]. Lauer et al., in their research work raised the issue of the criticality of the incubation period for COVID-19. They studied 181 confirmed cases and identified that the incubation period may vary from 5 days to 14 days and based on this better surveillance and control activities can be planned [14]. In recent research work, Singer analyzed data of 25 infected counties to follow short term predictions about the COVID 2019 outbreak. The research highlighted that the location-specific rate of disease spread follows either steady or explosive power-law growth with different scaling exponents. With this understanding, the authors analyzed the impact of lockdown in various parts of the world [15]. Based on the above literature, it is evident that sufficient work is available on exploratory data analysis to understand the existing trend of the epidemic but still there is a lot of scopes to develop and test efficient machine learning based prediction models so that proactive strategies could be identified to cater the immediate needs.

## 3 Dataset description

The day to day prevalence data of COVID-2019 from January 22, 2020, to April 1, 2020, were retrieved from the official repository of Johns Hopkins University. The dataset consists of daily case reports and daily time series summary tables. In the present study, we have taken time-series summary tables in CSV format having three tables for confirmed, death and recovered cases of COVID-2019 with six attributes i.e. province/state, country/region, last update, confirmed, death and recovered cases, where the update frequency of the dataset is once in a day [2]. Figure 5 presents the COVID-19 confirmed, recovered, and death cases distribution across the world since the time data was recorded. It is easy to observe the exponential growth of the spread which needs to be controlled.

**Fig. 5.**
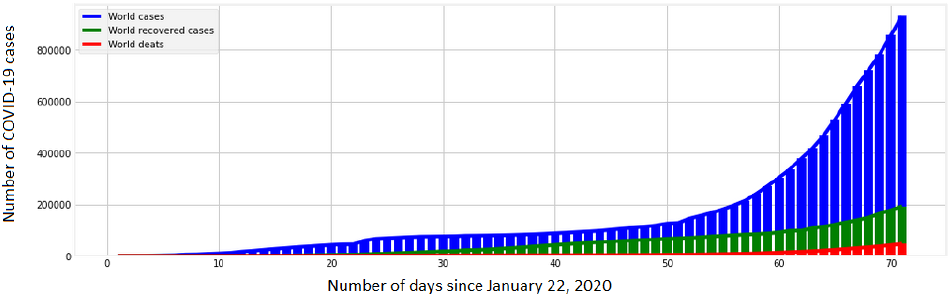
Distribution of COVID-19 cases across the globe.

## 4 Epidemic Analysis

The COVID-19 spread has brought the world under the brink of loss of human lives due to which it is of utmost importance to analyze the transmission growth at the earliest and forecast the forthcoming possibilities of the transmission. With this objective, state-of-the-art mathematical models are adopted based on machine learning such as support vector regression (SVR) [16] and polynomial regression (PR) [17], and deep learning regression models such as a standard deep neural network (DNN) and recurrent neural networks (RNN) using long short-term memory (LSTM) cells [18]. Machine learning and deep learning approaches are implemented using the python library “sklearn” and “keras” respectively, to predict the total number of confirmed, recovered, and death cases worldwide. The prediction will allow undertaking the necessary decisions based on transmission growth such as increasing the lockdown period, executing the sanitation procedure, providing the everyday resources, etc.

## 5 Training and testing

The regression approaches for epidemic analysis are trained and tested on realtime data [2] using the number of confirmed, recovered, and death cases as the label for the corresponding day. With extensive experiments, machine learning approaches are implemented with the polynomial kernel of degree 6 and other coefficient values as gamma=0.01, epsilon=1, and C=0.1. The standard DNN consists of a dense input layer with 128 neurons, three hidden dense layers with 256 neurons and output layer is consists of a single neuron whereas the RNN, consists of three stacks of LSTM layers having 64 neurons combined with 10% dropout to avoid the overfitting problem and final output layer with a single neuron. The mean squared error (MSE) is the most widely used objective function and root mean square error (RMSE) as a metric function for evaluating the regression models. The MSE loss can be computed by using equation 1.

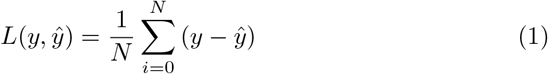

where y indicates the original value, 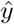 indicates the predicted value, and N is the number of samples predicted.

Due to the limited availability of the data, the trained models are validated against the training data and then utilized to forecast the number of confirmed, recovered, and death cases for the next 10 days.

## 6 Results and discussion

The discussed machine learning and deep learning approaches output the possible number of cases for the next 10 days across the world. Figure 6 illustrates the predicted trend of the COVID-19 using SVR, PR, DNN, and LSTM with worldwide data. Table 1 indicates the RMSE score of the approaches computed against the available number of COVID-19 cases. It is also observed that training of LSTM model is heavily dependent on the deviation in the values, with the fact that larger the deviation more the time it takes to train. Hence, the number of cases were scaled using minmax scaler to fit the LSTM model and later the predicted cases were rescaled to the original range using invert minmax transform from “sklearn” python library. Among these approaches, the visual representation of the prediction from figure 6 and RMSE score as highlighted in table 1 confirms the PR approach as the best fit to follow the growing trend.

**Fig. 6.**
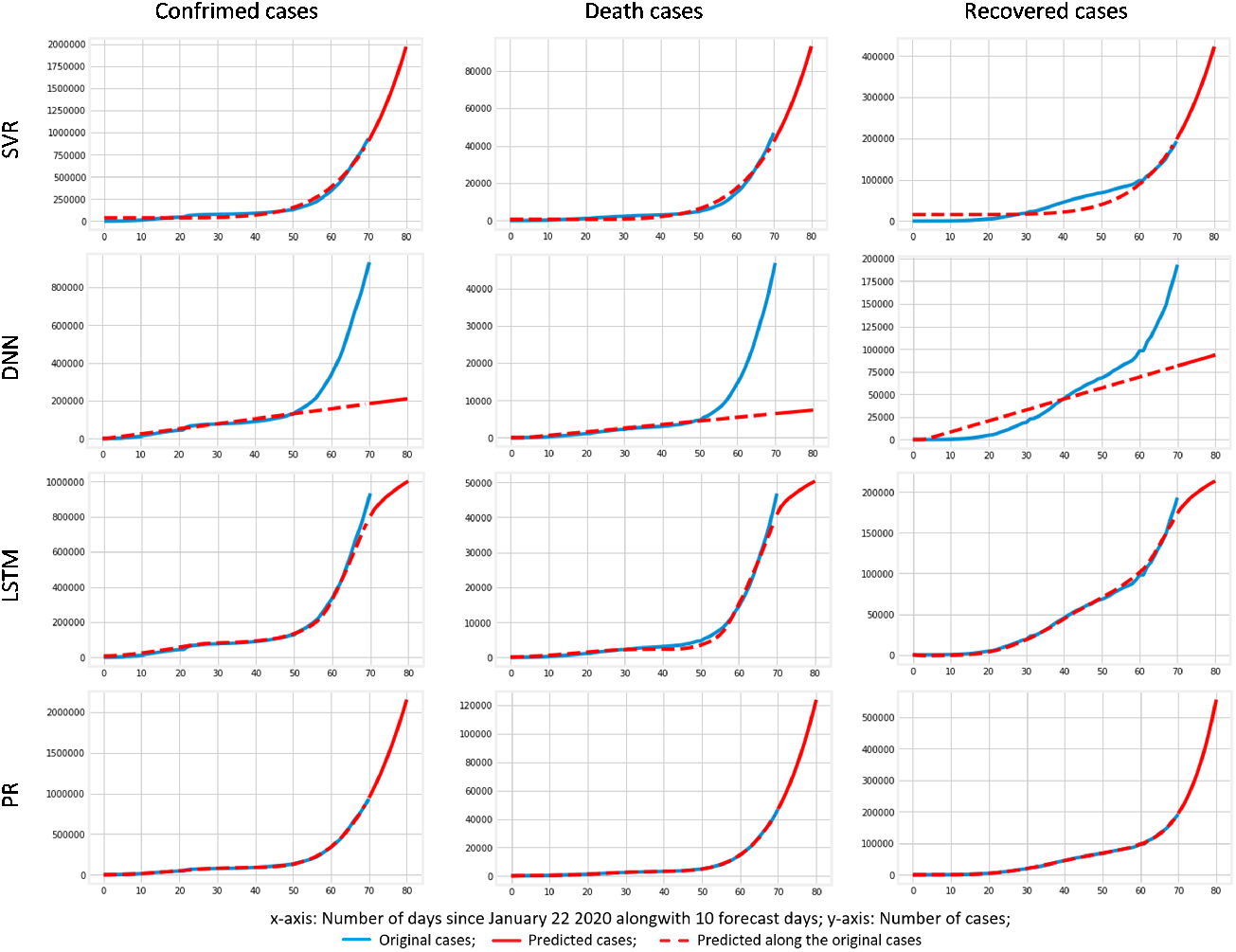
COVID-19 worldwide epidemic analysis using SVR, DNN, LSTM, and PR.

**Table 1.**
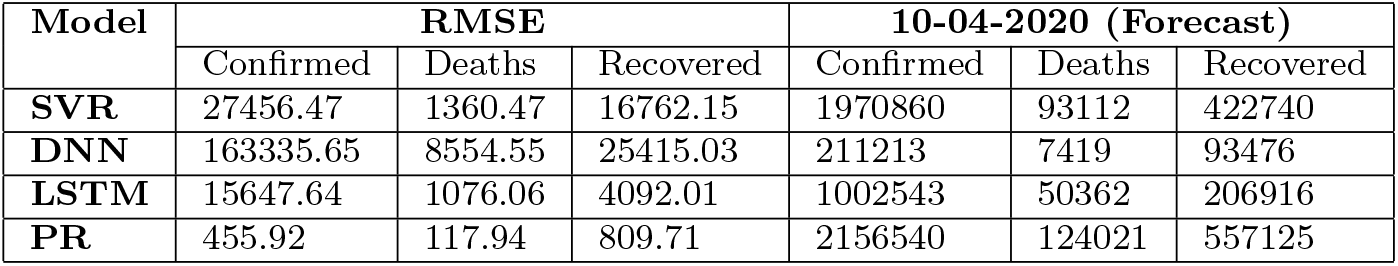
Comparative results.

## 7 Conclusion

The world is under the grasp of SARS-CoV2 (COVID-19) virus. Early prediction of the transmission can help to take necessary actions. This article proposed to utilize the machine learning and deep learning models for epidemic analysis using data from Johns Hopkins dashboard. The results show that polynomial regression (PR) yielded a minimum root mean square error (RMSE) score over other approaches in forecasting the COVID-19 transmission. However, if the spread follows the predicted trend of the PR model then it would lead to huge loss of lives as it presents the exponential growth of the transmission worldwide. As observed from China, this growth of the COVID-19 can be reduced and quenched by reducing the number of susceptible individuals from the infected individuals. This is achievable by becoming unsocial and following the lockdown initiative with discipline. The study can further be extended to utilize other machine learning and deep learning models.

## Data Availability

Novel Coronavirus COVID-19 (2019-nCoV) publicly available data repository by Johns Hopkins CSSE.

https://github.com/CSSEGISandData/COVID-19

## Compliance with Ethical Standards

### Ethical approval

This article does not contain any studies with human participants or animals performed by any of the authors.

## Acknowledgements

We are very much obliged to our institute, Indian Institute of Information Technology Allahabad (IIITA), India and Big Data Analytics (BDA) lab for ensuring the needed resources and support. Authors are also indebted to “ASEAN-India Science Technology Development Fund (AISTDF)”, SERB, Sanction letter no. – IMRC/AISTDF/R&D/P-6/2017, for their financial support to carry out the background research which helped significantly for the implementation of present research work.

